# Making the urban environment around schools healthier, developing an initial programme theory: an early realist process evaluation of the London superzones pilot

**DOI:** 10.1101/2020.02.04.20020396

**Authors:** Heather Catt, Steven L Senior

## Abstract

**BACKGROUND:** More than four out of five people in England live in an urban area so the urban environment is an important determinant of health and an important contributor to health inequalities. This is especially true for children, for whom the impacts of their local environment may have lifelong effects. To improve the health effects of the urban environment the London Devolution Partnership piloted multi-agency partnerships focusing on 400m ‘superzones’ around schools in deprived communities. The intention is for local partners to work together to make the environment within the superzone healthier.

**METHODS:** A realist process evaluation of 13 pilot sites. We used data from programme documents, a rapid literature search, and interviews with lead officers for the 13 local authorities that took part in the programme. Qualitative analysis was used to identify combinations of context, mechanisms, and outcomes (CMOs) that affected the creation of a local partnership and plan.

**RESULTS:** All but one of the 13 pilot sites established a superzone and multi-agency action plan. We identified 12 CMOs that affect the process of creating a school superzone. We present a logic model that describes how these CMOs combine to make up an initial programme theory of superzones. Key aspects of this programme theory include the importance of local partners’ priorities, leadership and support from across the local authority, and the flexibility of the model itself.

**CONCLUSION:** The superzones model is an emerging framework for local place-based partnership working to improve the urban environment. However, the success of the model is likely to depend on local context. The evidence presented here could guide the further roll out of the model, and support those local authorities that want to create superzones.

## INTRODUCTION

More than four out of five people in England live in an urban area [1]. Features of the urban environment that affect health and are potentially modifiable include air quality [2], density of unhealthy food outlets [3], infrastructure that supports active travel [4,5] and access to green spaces [6,7]. These features are distributed in a way that reinforces inequalities in health between more and less deprived areas. For example, more deprived urban areas tend to have worse air quality [8], more fast food outlets [9], and worse access to green spaces [10]. Further, these impacts may be felt most by children due to their narrower geographical horizons and the lifelong effects on health and wellbeing of childhood experiences [11]. Environments around schools affect children’s health through exposure to hazards such as poor air quality, fast food outlet density, smoking, alcohol and gaming/gambling [12–16].

The move of public health teams back to local authorities in April 2013 created an opportunity for local authorities to make more use of their powers to change the environment in a way that improves their population’s health. Six years on, progress appears mixed. While there are examples of guidance and good practice [17], research has found that three quarters of local plans do not reference the local authority’s health and wellbeing strategy or the health needs described in the local authority’s joint strategic needs assessment [18].

Devolution within England is creating regional areas led politically by elected mayors. While these regions vary in the extent to which they have taken control over health spending [19], they have the potential to act as a catalyst for changes in the urban environment that improve health [20]. In London, the Mayor’s Health Inequalities Strategy includes aims to make the environment healthier and to improve children’s health [21]. A London “Devolution Partnership” has been established with membership of Public Health England (PHE), the Greater London Authority (GLA), the NHS, London Councils and the Association of Directors of Public Health (ADPH).

The London Devolution Partnership piloted multi-agency partnerships focusing on 400m superzones around schools in deprived communities. By using a systems approach in partnerships, the superzones may be able to better exploit potential benefits and synergies while managing trade-offs, to allow for prioritisation of solutions to improve the health outcomes of children [22]. Thirteen pilot boroughs agreed to take part in the pilot and worked with local stakeholders to identify school superzones and develop a local model and action plan.

In consultation with the London devolution partners we conducted a process evaluation of the superzone pilots. The aim was to capture learning that took place during delivery, and in particular to identify the key factors that influenced the setting up of the superzone partnership and the creation of an agreed superzone action plan. We present an initial programme theory that describes how superzones work. The results provide evidence to inform the development of the 13 pilot sites, as well as any wider roll out and the theory can be further tested and refined with evaluation of the wider roll out of the superzones [23].

### Ethics

Ethical approval for the work was given by the University of Manchester (reference: 2019-6307-9575).

## METHODS

The school superzones concept was deliberately loosely defined and was intended to be adapted to meet local needs. To capture this variation in how the model worked across different pilot sites, we used a realist evaluation approach. Realist evaluation aims to identify what works, why, for whom and under what circumstances [24,25]. The method describes what happened using configurations of contexts, mechanisms and outcomes [23]. Mechanisms describe what is happening in the programme to bring about outcomes and the realist method considers in what contexts, “in what circumstances” and “for whom”, these mechanisms operate [24,25]. The method allows for explanation of why outcomes may vary across contexts and may be different to those that were originally anticipated [24,25]. These variations from the same intervention are captured in context-mechanism-outcome configurations (CMOCs) [24,25]. Together, the CMOCs make up a theory of the programme, which highlights the configurations needed for the interventions to work and why an interventions works in some areas and not others.

### Data collection

The evaluation used an exploratory, qualitative research design with data collected during May and June 2019. We used two methods of data collection: a review of documents from the superzones pilots and other literature on process evaluations of place-based programmes aimed at improving population health; and semi-structured interviews with the Public Health England (PHE) superzones project manager and the local authority coordinators of the 13 superzones.

#### Review of documents and literature on similar process evaluations

We obtained documents from the establishment of the superzones project (e.g. invitations to participate and briefing packs). We only included documents from the early establishment of the pilots so that they were uncontaminated by the learning as the project pilot developed. We coded these documents to identify an initial set of CMOs.

We sought evidence from the published research literature by doing a rapid literature search. Both researchers independently searched databases (MEDLINE, SCOPUS and PUBMED). Searches used a combination of terms (and their variants) including “partnership working”, “realist evaluation”, “process evaluation”, “pilot”, “place-based”, “urban”, “environment”, “health inequalities”, “air quality”, “child health” and “child obesity”.

#### Interviews with key stakeholders

We used purposive sampling to recruit participants in the superzone pilots. We interviewed the local authority staff with responsibility for establishing the superzones in each area to support the initial theory building [23]. The interviews were arranged by PHE and conducted over two days at PHE offices in London. Two researchers conducted interviews and alternated between interviewing and taking notes. Two interviewees were unable to attend on the day and were subsequently interviewed by phone using the same methods as the face to face interviews (ethical approval reference for this change: 2019-6307-11162). Thirteen interviews were done in total: twelve superzone leads and the PHE programme manager (one superzone lead was responsible for superzones in two local authorities).

The interviews were conducted prior to the document review to capture their views of the important contexts, mechanisms and outcomes of the process of establishing a superzone pilot and action plan. This was to avoid imposing our interpretation of the programme theory and reduce the risk of crowding out discussion of other important contexts, mechanisms and outcomes plan, and to avoid confirmation bias [26]. However, we analysed the documents prior to the analysis of the interview data to ensure that we tested the initial programme theory against the views of the pilot participants. We used a semi-structured topic guide that included questions on the role of the individual in the pilot, who else they involved, how they developed an action plan, as well as the initial motivations and challenges or enablers the project faced. The interview process took place in May to July 2019.

### Data Analysis

The documents were analysed using the process described by Bowen and applied by Mukumbang et al [27,28]. The data from the documents were coded as context, mechanism or outcome, according to the definitions within realist methodology [24,25]. Thematic framework analysis was used to identify common themes and to specify initial context-mechanism-outcome configurations.

The interview transcripts were coded as context, mechanism or outcome and the data collected was used to test and refine the CMO configurations from the document analysis. The resulting theories were reported back to the interview participants in a focus group to test their validity and further iterate the theory [23].

The results are reported in line with the realist evaluation reporting standards, RAMESES II [29].

## RESULTS

### Review of documents and literature search

Table 1 shows the types and descriptions of the four documents included in the document review. The literature searches did not identify any reports of similar process evaluations which could complement the theory developed from the policy documents.

**Table 1:** A description of the documents included in the document review.

| Title | Author / date | Description |
| --- | --- | --- |
| Seeking expressions of interest - London school health superzones pilot | London devolution partners, May 2018 | A letter sent to all London local authorities introducing them to the superzones concept and inviting them to express an interest in participating. |
| London devolution school superzones pilot project - Invitation to participation | London devolution partners, July 2018 | A letter sent to London local authorities following their expression of interest. The letter outlines the conditions to be able to join the pilot, the background and aims of the pilot and the support available to pilot authorities. |
| School superzones pilot project local authority briefing pack | London devolution partners, July 2018 | The briefing pack outlines the superzones initiative, its links to devolution, how it could contribute to improve child health and wellbeing and how local authorities can participate. It also highlights the potential related funding opportunities, timelines of the pilot and the mapping resources available. |
| Guide for school superzones pilot | London devolution partners, August 2018 | This document outlines a stepped approach to establishing a superzone. It provides deadlines, details of linked projects and sources of support for local authorities involved in the pilot. |

### Outcomes

All but one of the pilot areas established local partnerships and developed a superzone action plan. Of those that did, all felt that creating the superzone had helped to improve partnership working either by building new relationships or strengthening existing ones by giving them a focus. As this is a process evaluation, it is too early to draw conclusions about the health impacts of the superzones. For evaluation purposes, the outcome was the creation of an agreed action plan and governance structure for the superzone.

### An initial programme theory of school superzones

The review of programme documents identified 19 distinct context-mechanism-outcome configurations (CMOCs). These were further refined following analysis of the interview transcripts, leaving a final set of 12 CMOCs (see Table 2 below). Together these make up an initial programme theory of school superzones, which is shown in Figure 1 and described below.

**Table 2:**
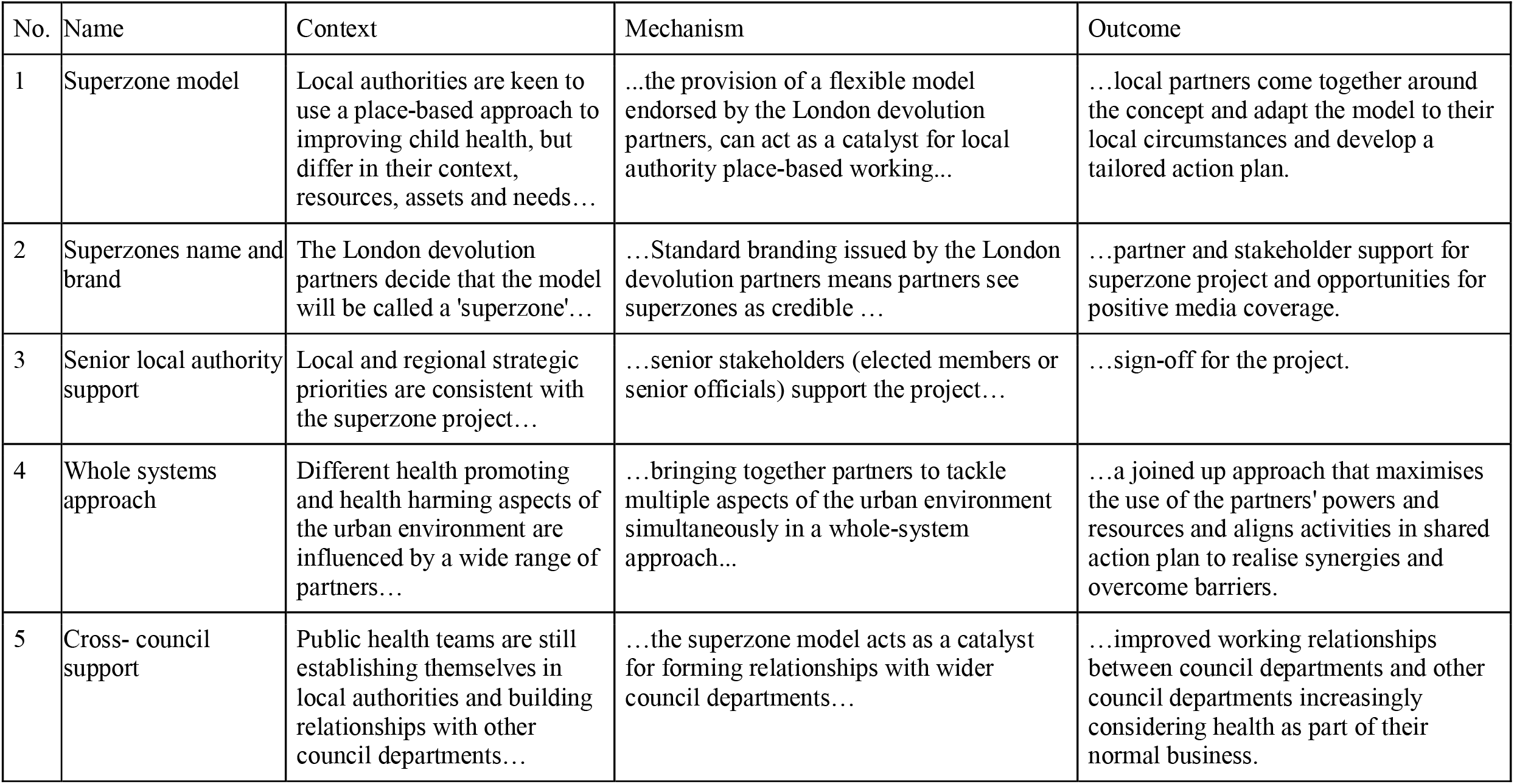
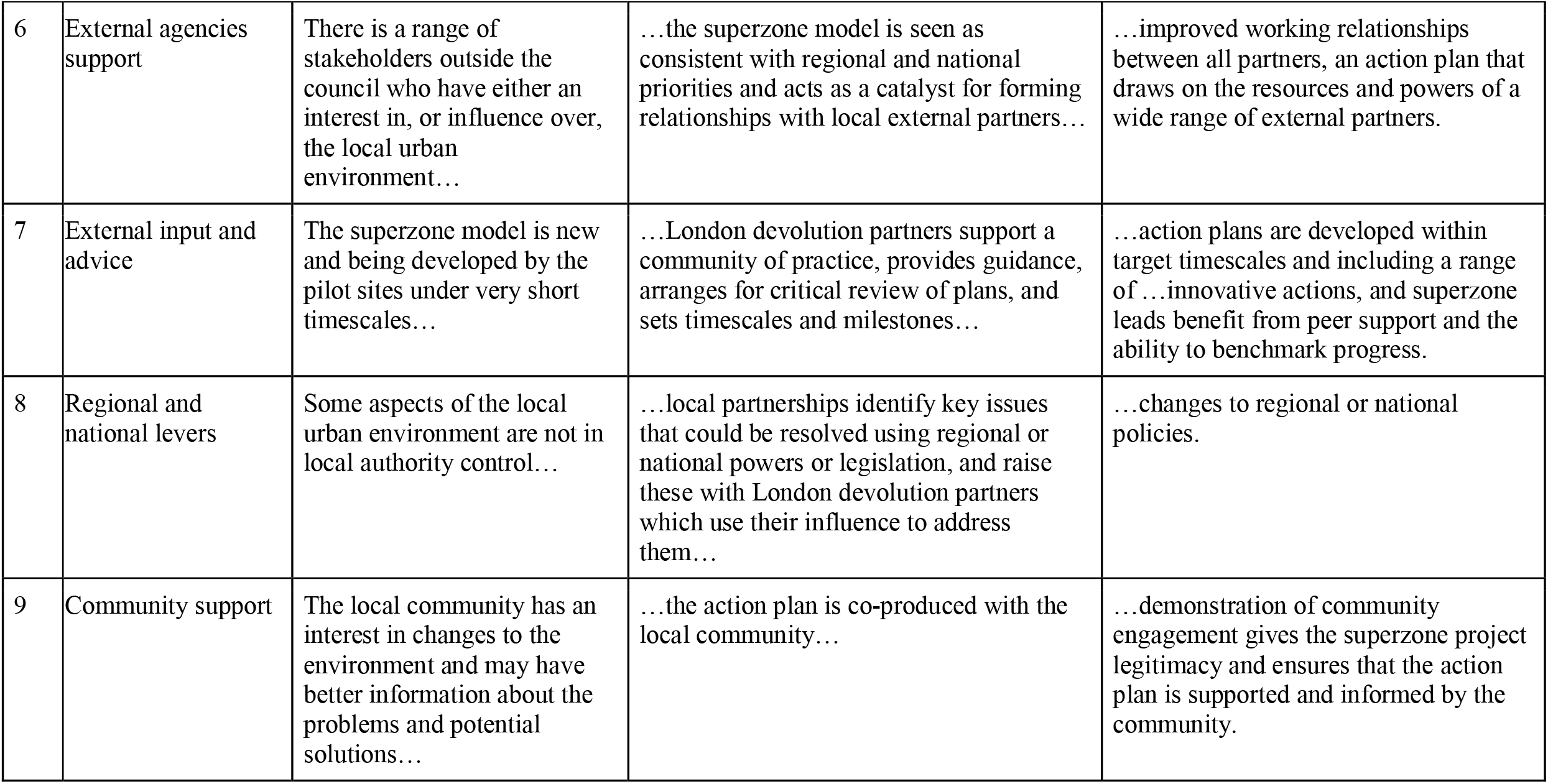
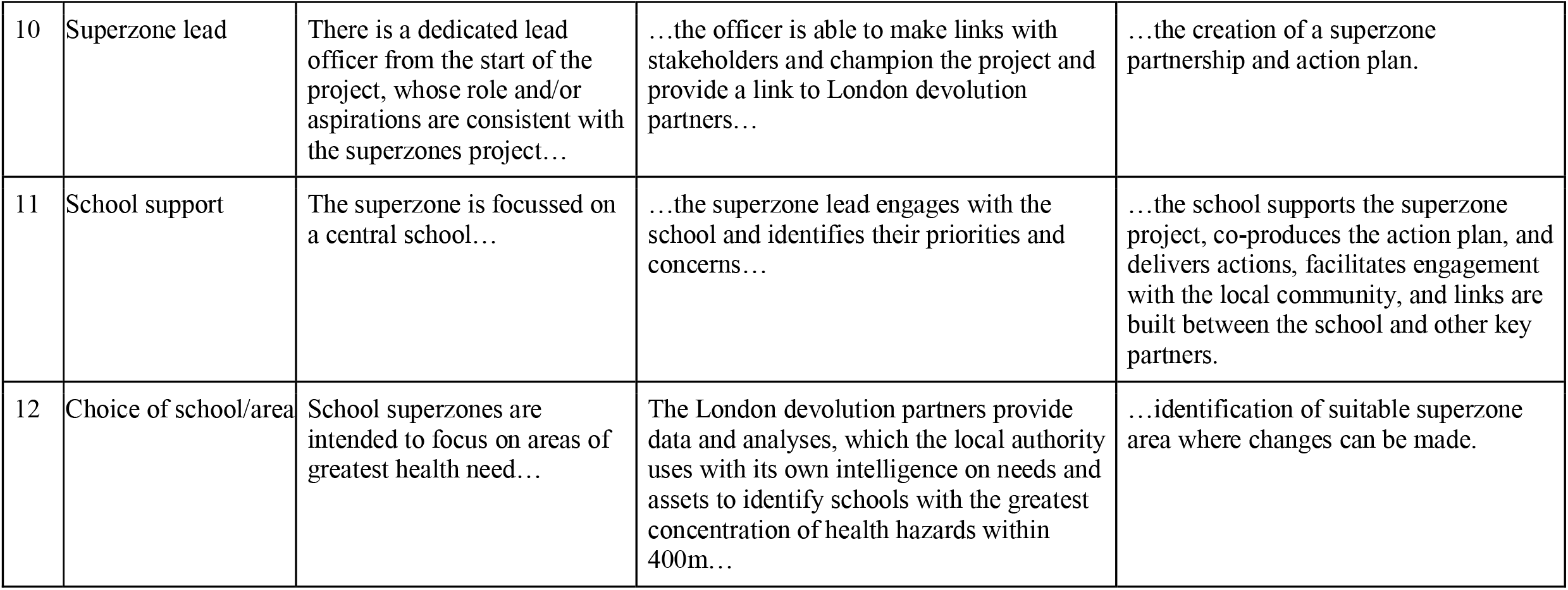
Context-Mechanism-Outcome configurations.

| No. | Name | Context | Mechanism | Outcome |
| --- | --- | --- | --- | --- |
| 1 | Superzone model | Local authorities are keen to use a place-based approach to improving child health, but differ in their context, resources, assets and needs... | ...the provision of a flexible model endorsed by the London devolution partners, can act as a catalyst for local authority place-based working... | ...local partners come together around the concept and adapt the model to their local circumstances and develop a tailored action plan. |
| 2 | Superzones name and brand | The London devolution partners decide that the model will be called a 'superzone'... | ...Standard branding issued by the London devolution partners means partners see superzones as credible ... | ...partner and stakeholder support for superzone project and opportunities for positive media coverage. |
| 3 | Senior local authority support | Local and regional strategic priorities are consistent with the superzone project... | ...senior stakeholders (elected members or senior officials) support the project... | ...sign-off for the project. |
| 4 | Whole systems approach | Different health promoting and health harming aspects of the urban environment are influenced by a wide range of partners... | ...bringing together partners to tackle multiple aspects of the urban environment simultaneously in a whole-system approach... | ...a joined up approach that maximises the use of the partners' powers and resources and aligns activities in shared action plan to realise synergies and overcome barriers. |
| 5 | Cross- council support | Public health teams are still establishing themselves in local authorities and building relationships with other council departments... | ...the superzone model acts as a catalyst for forming relationships with wider council departments... | ...improved working relationships between council departments and other council departments increasingly considering health as part of their normal business. |
| 6 | External agencies support | There is a range of stakeholders outside the council who have either an interest in, or influence over, the local urban environment... | ...the superzone model is seen as consistent with regional and national priorities and acts as a catalyst for forming relationships with local external partners... | ...improved working relationships between all partners, an action plan that draws on the resources and powers of a wide range of external partners. |
| 7 | External input and advice | The superzone model is new and being developed by the pilot sites under very short timescales... | ...London devolution partners support a community of practice, provides guidance, arranges for critical review of plans, and sets timescales and milestones... | ...action plans are developed within target timescales and including a range of ...innovative actions, and superzone leads benefit from peer support and the ability to benchmark progress. |
| 8 | Regional and national levers | Some aspects of the local urban environment are not in local authority control... | ...local partnerships identify key issues that could be resolved using regional or national powers or legislation, and raise these with London devolution partners which use their influence to address them... | ...changes to regional or national policies. |
| 9 | Community support | The local community has an interest in changes to the environment and may have better information about the problems and potential solutions... | ...the action plan is co-produced with the local community... | ...demonstration of community engagement gives the superzone project legitimacy and ensures that the action plan is supported and informed by the community. |
| 10 | Superzone lead | There is a dedicated lead officer from the start of the project, whose role and/or aspirations are consistent with the superzones project... | ...the officer is able to make links with stakeholders and champion the project and provide a link to London devolution partners... | ...the creation of a superzone partnership and action plan. |
| 11 | School support | The superzone is focussed on a central school... | ...the superzone lead engages with the school and identifies their priorities and concerns... | ...the school supports the superzone project, co-produces the action plan, and delivers actions, facilitates engagement with the local community, and links are built between the school and other key partners. |
| 12 | Choice of school/area | School superzones are intended to focus on areas of greatest health need... | The London devolution partners provide data and analyses, which the local authority uses with its own intelligence on needs and assets to identify schools with the greatest concentration of health hazards within 400m... | ...identification of suitable superzone area where changes can be made. |

**Table 3:** Table of CMOCs following analysis of interviews.

| No. | Label | Context | Mechanism | Outcome |
| --- | --- | --- | --- | --- |
| 1 | Senior local authority support | Local and regional strategic priorities are consistent with the superzone project... | ...senior stakeholders (elected members or senior officials) support the project... | ...sign-off for the project. |
| 1a | Senior local authority support | There is doubt about whether the superzone project fits with local priorities, or the ability to achieve anything without funding... | ...the superzone lead must be able to convince senior stakeholders of the benefits of the project... | ...sign-off for the project. |
| 1b | Senior local authority support | There is a change of key senior stakeholder (e.g. cabinet portfolio holder or DPH)... | ...the senior stakeholder is no longer committed to the superzone project... | ...absence of support for commitment of public health team or other council resource to the superzone project. |
| 1c | Senior local authority support | There is an elected member who champions the project (either the relevant ward councillor or portfolio holder)... | ...other council departments see the superzone project as more of a priority... | ...enhanced support from other council departments. |
| 1d | Senior local authority support | The timing of the superzones project aligns (or does not align) with the local authority budgeting or priority setting cycle... | ...the superzone project is (is not) incorporated into wider local authority strategies, plans, or budgets... | ...enhanced (or diminished) support from across the council. |
| 2 | Superzone model | Local authorities are keen to use a place-based approach to improving child health, but differ in their context, resources, assets and needs... | ...the provision of a flexible model targeting changes to the environment within a 400m radius outside of the school, endorsed by the London devolution partners, can act as a catalyst for local authority place-based working... | ...local partners come together around the concept and adapt the model to their local circumstances and develop a tailored action plan. |
| 2a | Superzone model | There is already a mature partnership working between public health and planning departments involving working together on another strategic approach to healthy places... | ...the superzone pilot may be seen as a backwards step... | ...failure to engage with the superzone pilot. |
| 2b | Superzone model | The local authority experiences uniformly high levels of deprivation... | ...a focus on a single school zone may be seen as inappropriate and the creation of an extra administrative zone may confuse stakeholders... | ...failure to engage with the superzone pilot. |
| 2c | Superzone model | The 400m radius excludes key assets or is not practical in a small local authority area or conflicts with more relevant administrative boundaries... | ...the partners may adapt the core features of the model... | ...achieving support from local partners and stakeholders and/or taking advantage of other opportunities. |
| 2d | Superzone model | There is other complementary work happening in the school... | ...the inclusion of in-school actions in the superzone feels that the superzone project is supporting its priorities... | ...enhanced engagement with the school. |
| 2e | Superzone model | Local stakeholders have priorities that are not included in the suggestions from London devolution partners... | ...the inclusion of local partners' priorities helps partners to feel that the superzone is addressing their needs or concerns... | ...greater support from local partners for the superzone project. |
| 2f | Superzone model | There is experience in the public health team of cross-council working... | ...the superzone model provides a template for place-based working... | ...focusing existing partnership working on the superzone. |
| 2g | Superzone model | There are other current or planned projects or other initiatives either in the superzone area, or across the local authority... | ...the incorporation of these other projects or initiatives into the superzone action plan... | ...extra resources to support the superzone aims and greater partnership support. |
| 3 | Whole systems approach | Different health promoting and health harming aspects of the urban environment are influenced by a wide range of partners... | ...bringing together partners to tackle multiple aspects of the urban environment simultaneously in a whole-system approach... | ...a joined up approach that maximises the use of the partners' powers and resources and aligns activities in shared action plan to realise synergies and overcome barriers. |
| 3a | Whole systems approach | There a very mature level of whole-systems working... | ...partners may see little benefit in the superzone model... | ...more limited engagement with the superzone model. |
| 3b | Whole systems approach | There are existing wider initiatives or ongoing regeneration projects or available development land... | ...the superzone focuses these wider activities into the same area... | ...to benefit from resources and synergies between apparently different projects and the ability to influence regeneration. |
| 3c | Whole systems approach | There are limited opportunities to make meaningful changes to the physical environment... | ...partners may lose enthusiasm for the superzone project... | ...limited engagement or loss of momentum of the project. |
| 3d | Whole systems approach | Public health is not reflected in the local plan or planning not reflected in health and wellbeing strategy or joint strategic needs assessment... | ...planning powers are not available to support the superzone actions... | ...failure to use local planning levers to achieve change. |
| 3e | Whole systems approach | National legislation or regional policy does not enable the use of local fiscal or policy levers for the purpose of promoting health... | ...local authority departments feel unable to test these powers as part of the school superzone... | ...failure to fully exploit the powers available to the local authority. |
| 4 | Cross-council support | Public health teams are still establishing themselves in local authorities and building relationships with other council departments... | ...the superzone model acts as a catalyst for forming relationships with wider council departments... | ...improved working relationships between council departments, other council departments increasingly considering health as part of their normal business, and commitment of other council departments to actions in the superzone action plan. |
| 4a | Cross-council support | There are already strong working relationships within the council... | ...the public health team is able to establish strong governance and cross-council engagement, taking account of other department's priorities and needs... | ...increased ownership of actions in the action plan by other council departments. |
| 4b | Cross-council support | There is visible support from elected members or senior council officers or there is a fit between the superzone project and wider departments' priorities... | ...other council departments see the superzone project as more of a priority... | ...increased engagement of other council departments with the superzone project. |
| 4c | Cross-council support | There is no funding to support the superzone project... | ...Lack of funding or short timescales causes scepticism among local partners about the viability of the superzone project... | ...more limited engagement with the superzone project. |
| 5 | External agencies support | There is a range of stakeholders outside the council who have either an interest in or influence over the local urban environment... | ...the superzone model is seen as consistent with regional and national priorities and acts as a catalyst for forming relationships with local external partners... | ...improved working relationships between all partners, the action plan draws on the resources and powers of a wide range of external partner, and external partners feel that their priorities are being met through the superzone project. |
| 5a | External agencies support | There many pre-existing projects in the same space... | ...the external partners may suffer from intervention fatigue or be confused about how the superzone fits with these other projects... | ...reduced engagement of external partner with the superzone project. |
| 5b | External agencies support | There is no funding to support the superzone project... | ...Lack of funding or short timescales causes scepticism among local partners about the viability of the superzone project... | ...more limited engagement from external partners with the superzone project. |
| 6 | External input and advice | The superzone model is new and being developed by the pilot sites under very short timescales... | ...London devolution partners support a community of practice, provides guidance, arranges for critical review of plans, and sets timescales and milestones... | ...action plans are developed within target timescales and including a range of innovative actions, solutions to shared barriers are identified, and superzone leads benefit from peer support and the ability to benchmark progress. |
| 6a | External input and advice | Local authority public health team has limited capacity and/or other competing priorities... | ...superzone leads may struggle to engage with support available... | ...failure to benefit from external advice and guidance. |
| 6b | External input and advice | The review of action plans is descriptive rather than critical... | ...superzone leads may not be able to judge whether they are making adequate progress... | ...limited impact on the development of action plans. |
| 6c | External input and advice | There is limited time for the superzone project and there is uncertainty about the future of the project beyond the pilot phase... | ...partners may see the timescales as unrealistic... | ...reduced engagement of partners with the superzone project. |
| 6d | External input and advice | Partners have competing priorities and pressures... | ...London devolution partners sets deadlines, sends reminders, and requests progress updates, which the superzone lead can use to maintain momentum... | ...action plans being developed within target timescales. |
| 6e | External input and advice | The local authority public health team already has experience of working closely with the planning department... | ...the local authority may see little benefit in the planning workshop... | ...little change in action plans. |
| 6f | External input and advice | The superzone model is new and being developed by the pilot sites under very short timescales... | ...superzone leads felt supported by a group of peers facing the same challenges... | ...a desire for the community of practice to continue beyond the pilot phase. |
| 7 | Regional and national levers | Some aspects of the local urban environment are not in local authority control... | ...Local partnerships identify key issues that could be resolved using regional or national powers or legislation and raise these with London devolution partners which uses its influence to change them... | ...changes to regional or national policies. |
| 7a | Regional and national levers | Changes to regional and national policies take time and political capital and the pilot is operating on a short timescale... | ...Local partnerships have been led to expect change quicker than is feasible... | ...a perception that there has been little change to regional and national policies and legislation. |
| 8 | Community support | The local community has an interest in changes to the environment and may have better information about the problems and potential solutions... | ...the action plan is co-produced with the local community... | ...demonstration of community engagement gives the superzone project legitimacy and ensures that the action plan is supported and informed by the community. |
| 8a | Community support | The local community has a high prevalence of low paid shift work or zero-hours contracts or limited English language ability... | ...the partnership may be unable to engage with the local community... | ...lack of involvement of community in developing the superzone action plan, consequent loss of legitimacy, and possible community resistance to actions. |
| 8b | Community support | The focal school is not engaged with the superzone partnership... | ...the school will not be able to act as a broker between the partnership and the local community... | ...lack of involvement of community in developing the superzone action plan, consequent loss of legitimacy, and possible community resistance to actions. |
| 8c | Community support | Previous community engagement or insight work has been done... | ...the community may experience consultation fatigue, and/or the local authority may have good understanding of community priorities... | ...reduced need for community engagement as part of the superzone project. |
| 8d | Community support | The superzone project is a pilot of a partially defined model with limited time and funding, and relationships between community and partners is limited... | ...Council officers may be wary of 'dipping in and out' of a community and raising expectations... | ...lack of involvement of community in developing the superzone action plan, consequent loss of legitimacy, and possible community resistance to actions. |
| 8e | Community support | It is possible to achieve 'quick wins'... | ...the community can see that the superzone is achieving things that matter to them... | ...enhanced community engagement with and support for the superzone project. |
| 9 | Superzone lead | There is a dedicated lead officer from the start of the project, whose role and/or aspirations are consistent with the superzones project... | ...the officer is able to make links with stakeholders and champion the project and provide a link to London devolution partners and other regional stakeholders... | ...the creation of a superzone partnership and action plan. |
| 9a | Superzone lead | There is a lack of capacity in the public health team, the lead officer has many competing priorities and limited time to commit to the superzone... | ...the ability to build a wider partnership is limited and the superzone has limited ability to take part in external events... | ...a more limited local partnership, action plan, less progress, and the superzone lead may not benefit from peer support network. |
| 9b | Superzone lead | The lead officer is already connected with local partners e.g. through wider role or previous work... | ...there will be existing personal relationships with important superzone partners... | ...enhanced stakeholder and/or partner support for the superzone project and action plan. |
| 9c | Superzone lead | There is a change of superzone lead after the start of the project... | ...personal relationships and links with stakeholders need to be rebuilt... | ...potential delay in achieving action plan or progress against plan. |
| 9d | Superzone lead | There is extra support for the superzone lead... | ...the ability to consistently build and maintain relationships with partners, and to project manage the superzone is enhanced... | ...more momentum and interest from partners in the superzone project. |
| 9e | Superzone lead | The superzone lead is a relatively senior officer in the council (e.g. consultant in public health) | ...the superzone lead is more able to influence senior stakeholders to support the project... | ...enhanced senior buy-in for the project. |
| 10 | School support | The superzone is focussed on a central school... | ...the superzone lead engages with the school and identifies their priorities and concerns... | ...the school supports the superzone project, co-produces the action plan, and delivers actions, facilitates engagement with the local community, and links are built between the school and other key partners. |
| 10a | School support | Relationships between either the school and the local authority education department are weak (e.g. because the school is part of an academy trust); or the relationship between the local authority public health and education department is weak and the public health team does not have a direct relationship with the school... | ...engagement with the school will be limited... | ...lack of support from the school, failure to co-produce the action plan with the school, greater difficulty engaging the local community, and lack of key information (e.g. travel plans). |
| 10b | School support | There are many other recent local initiatives or consultations... | ...the school or the local authority education department may feel that the superzone represents too great an additional burden on the school... | ...limited engagement from the school and potential damage to relationships between school, education department, and public health department. |
| 10c | School support | There is no funding to support the superzone project and limited pilot timescales and no clarity on the future of the project... | ...the superzone team may feel that they have nothing to offer the school and worry about raising expectations... | ...not engaging with the school |
| 10d | School support | The school has other competing pressures (e.g. Ofsted inspection, exams, or other initiatives) | .....a mechanism for change is: the school may be unable or unwilling to engage with the superzone project... | ...limited or variable engagement from the school. |
| 11 | Choice of school/area | School superzones are intended to focus on areas of greatest health need... | .....a mechanism for change is: the London devolution partners provide data and analyses, which the local authority uses with its own intelligence on needs and assets to identify schools with the greatest concentration of health hazards within 400m... | ...identification of suitable superzone area where changes can be made. |
| 11a | Choice of school/area | There is additional insight into local stakeholder interests (including the school, ward councillor), and likely levels of engagement and resources among key partners... | ...a mechanism for change is: the prioritisation process draws in wider context around the school and involves other stakeholders (e.g. local politicians)... | ...identification of an area with a combination of both high need and opportunity, the superzone project is more likely to get support from key stakeholders and partners. |
| 11b | Choice of school/area | There is no additional insight into whether key challenges are within local control... | .....a mechanism for change is: the prioritisation process does not take account of the feasibility of changing the key features of the chosen area... | ...superzone unable to address the key health hazards in its area. |
| 12 | Superzones name and brand | London devolution partners decide that the model will be called a 'superzone'... | ...standard branding issued by the London devolution partners means stakeholders see superzones as credible ... | ...partner and stakeholder support for superzone project and opportunities for positive media coverage. |
| 12a | Superzones name and brand | There are lots of other funded projects and opportunities... | ...'Superzone' term seen as unhelpful... | ...superzone term not being used and missed opportunities for positive media coverage. |
| 12b | Superzones name and brand | Council partners have engaged enthusiastically with the superzone model but external local partners may or may not... | ...standard 'superzone' branding across all pilot sites means stakeholders see the superzone project as part of a wider initiative and the local superzone gains credibility... | ...better engagement with non-council local partners (e.g. schools) and promotion of the achievements of the superzone partnership. |
| 13 | Measuring and demonstrating progress | Local authority analytical capacity is available and baseline data is available or can be collected... | ...the superzone partnership can collect and use data on outputs and outcomes to show partners that progress is being made... | ...continued support from partners for the project and support from senior stakeholder for continuing beyond the pilot phase. |

**Figure 1:**
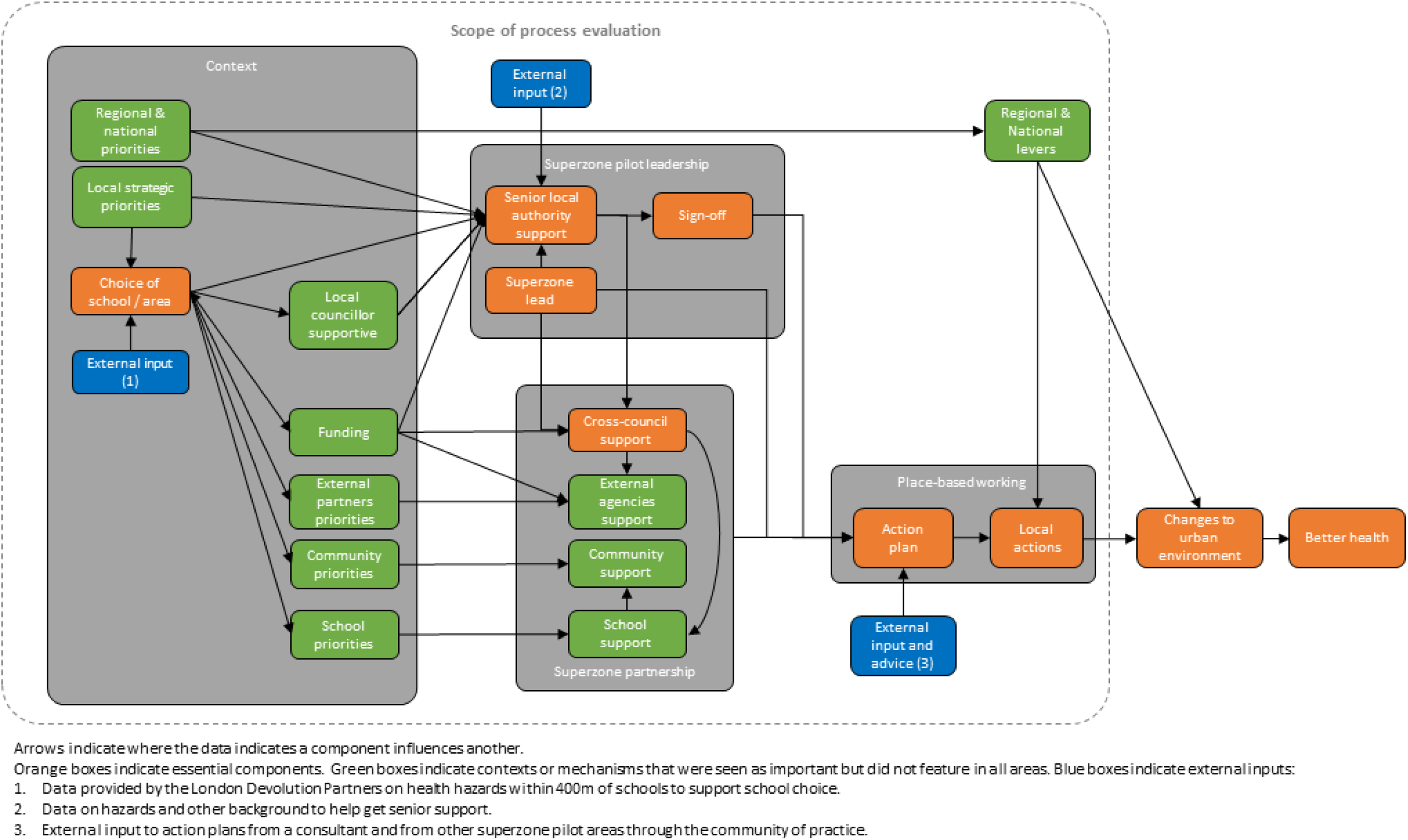
London Superzones Initial Programme Theory Flowchart.

**Figure 2:**
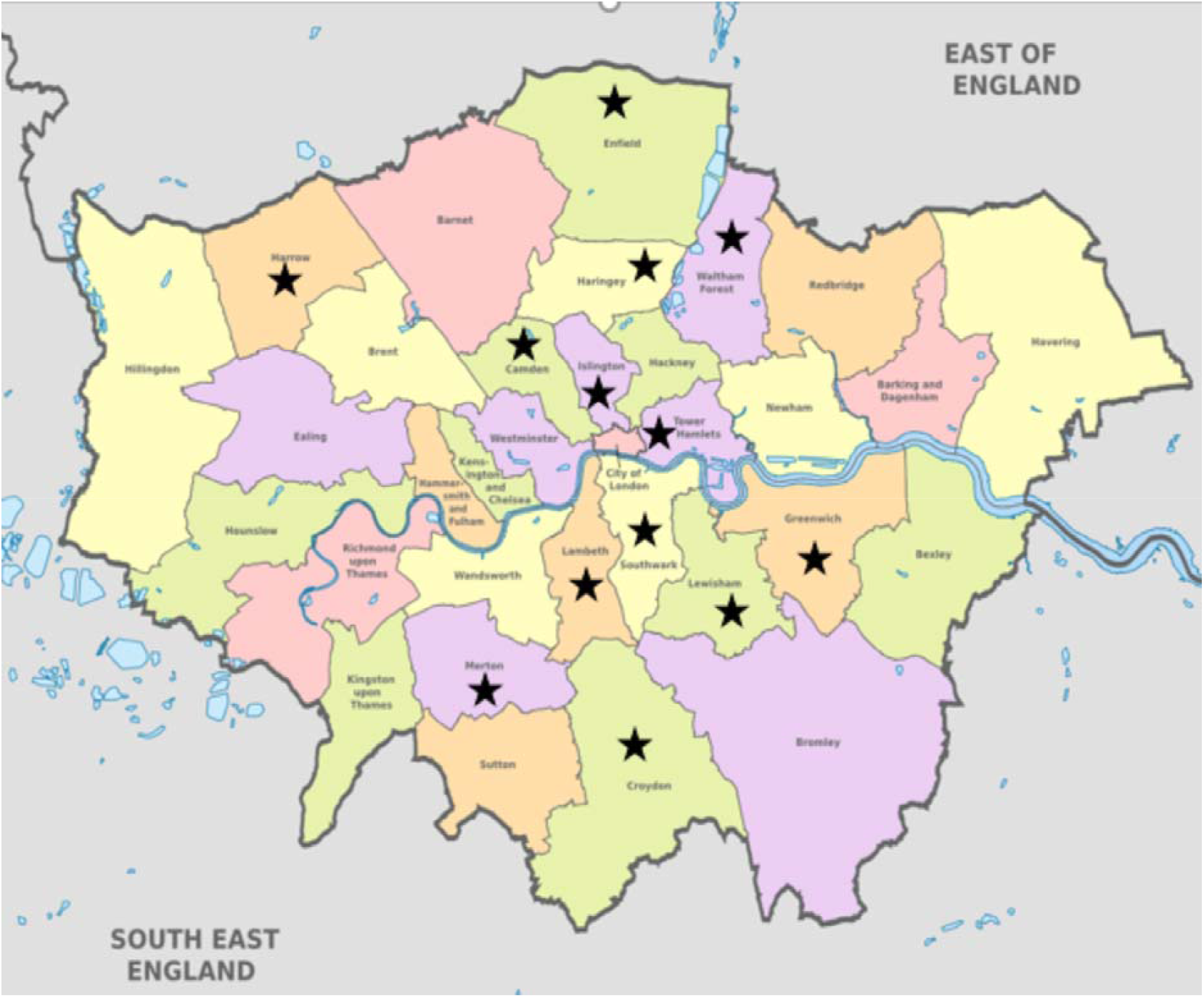
Map of local authorities who took part in the London superzones pilot.

### The superzone model

The superzone model emerged as the overarching CMOC. Interview participants reported that the superzone model, using a place-based approach to change the environment in a 400m radius around a central school, acted as a catalyst for partnerships to agree a set of shared goals and actions.

We identified additional contexts in which the mechanism either did not operate, or operated differently. For example, in areas with existing extensive work on planning and health, the superzone model was viewed as a backward step by key partners in the council, which meant that the mechanism did not operate, and a superzone was not set up. Similarly, in areas experiencing uniformly high levels of deprivation, the selection of a single zone to focus on was not acceptable to key partners, and the mechanism did not operate.

The flexibility of the superzone model was important and many areas reported that they varied aspects of the model to enable the central mechanism of partnership working to operate. Examples included changing the geographic boundary to include key local assets, or including in-school actions in the plan to help to engage the school as part of the partnership.

### Superzone name and brand

The project was designed using the term “superzone” and branding materials with the superzone project partners’ logos to add credibility and hopefully lead to partner support. However, in areas where there were already lots of branded partnership projects, the addition of another one was seen as unhelpful and in some cases the brand and name were not used in delivery of the pilot. Conversely, some areas with less existing partnership working felt that more branded materials would have strengthened the credibility of the project and helped to engage wider partners.

### Implementing the superzone model

As illustrated in Figure 1, the initial programme theory included four essential components that make up the core of the superzone implementation:

1. Choice of school/area
2. Superzone pilot leadership;
3. Superzone partnership; and
4. Place-based working.

#### Choice of school/area

A key CMOC was that externally provided data and support (identified as external input (1) in figure 1) was used to identify areas with the greatest level of need where changes can be made. However, without the addition of local intelligence on stakeholder priorities and levels of engagement and resources with partners, it was possible that areas or schools were selected where the challenges were not within local control or actions did not have political or community support. This prevented the mechanism from operating.

#### Superzone leadership

Leadership for the superzone pilots was essential, both in terms of senior leadership approving and supporting the superzone pilot, and project leadership to build and maintain partnerships and oversee the development of the action plan.

An original CMOC from the project documents was that project sign off would be given by senior leaders as the superzone pilot aligned with local and regional strategic priorities. The interviews highlighted that a strong superzone lead may be needed to convince senior leaders of the alignment and obtain sign off. Contexts that could disrupt this CMO include a change in senior leadership which weakens the council commitment to the pilot, or conversely a senior champion who strengthens the council commitment. Either one would see a change in the resources and staff committed to the pilot. The alignment (or otherwise) of the superzones pilot with budgeting or planning cycles could also affect the CMOC and result in sign off of the pilot but limited commitment of resources.

The nomination of a dedicated lead officer to project manage the superzone pilot was also a prerequisite for participation in the pilot programme. By linking with stakeholders and the London devolution partners, the dedicated lead would be able to ensure the superzone partnership and action plan were delivered. In interviews, the superzone leads reported that the pilots had required significant time commitment to build and maintain relationships across the council and with wider partners. Several mentioned this as a barrier to scaling up the projects in future. Several contexts were identified that affected this CMOC. Increased seniority of the lead, the degree of ‘fit’ with the lead’s wider portfolio of work and the availability of capacity within the wider public health team to support the lead, all increased the amount of time that the lead could commit to the project. As with the level of senior support for the pilot, the ability of the superzone lead to coordinate the project could be eroded by a change of staff or priorities.

#### Superzone partnership

Participants in the interviews talked about four distinct partnerships that made up the superzone: the partnership between council departments; partnerships with external agencies (such as housing associations or local voluntary sector organisations); the partnership with the school; and the partnership with the local community. Of these four, only the cross-council partnership was essential.

##### Cross-council support

Partnership working between the public health team and wider council departments was at the core of all the superzone pilots. Where public health teams were still establishing relationships across the local authority, the superzone project acted as a catalyst for forming relationships, enabling partnership working and increasing the consideration of health as part of the core business of all departments. In the context of a superzone built on a history of previous partnership projects, this mechanism was reinforced leading to increased ownership of the action plan. The involvement of a senior council officer or elected member as a champion for the superzone pilot also improved engagement of wider council departments. In some areas the lack of funding and short timescales created scepticism amongst partners and limited their engagement.

##### External agencies’ support

In a context where aspects of the urban environment are under the influence of agencies outside the council, the superzone model intended to act as a catalyst to wider partnership working. Examples of the types of agencies who were involved in the superzone pilots include: housing associations, local charities, police, and local businesses, although some areas did not work with external agencies. The lack of funding attached to the pilot was a barrier to engagement of external partners in some areas, as was the existence of many pre-existing projects in the same space.

Conversely, in contexts where external partners had relevant powers of funding, the pilot enabled the local authority to establish connections that had previously not existed.

##### School support

In its guidance, the London devolution partners highlighted engaging the schools in the superzone as an important mechanism. Almost all superzone pilots tried to involve the school in the superzone project. Success was mixed. Where schools were engaged, reported outcomes included the school acting as a gateway to engaging the local community and building relationships between the school and other key partners. Contexts that resulted in limited or no school engagement included where relationships between the education department and public health or the school were weak (making it difficult to build relationships with the school), where schools had academy trust status, or competing priorities, such as upcoming Ofsted inspections or exams or where there were other local projects or consultations that had created fatigue. This did not stop areas developing a superzone and an action plan, but they did not benefit from the school’s support or its links to the local community.

##### Community support

The CMOC for community support suggests that co-production of the plan with the community would bring both legitimacy and insight into the challenges in an area. Some pilot areas put substantial effort into engaging with the local community. The response varied depending on existing local issues. Where council officers viewed the superzones model in terms of its limitations of time and budget, there was a reluctance to be seen to “dip in and out” of the community and raise expectations. Further contexts which restricted the CMOC included consultation fatigue arising from previous projects, or difficult relationships between the school and the community, and a high prevalence of shift workers, zero hours contracts or people for whom English was an additional language. The inclusion of quick win activities to meet the concerns of the community could strengthen community engagement.

#### Place-based working

The original intention was that the superzones would become a focus for both existing and new actions, and that by bringing these together in a single place-based action plan synergies between actions would lead to the total improvement in the area being greater than the sum of the individual actions. Contexts in which the mechanism would not operate included where there were mature partnership working already, where there are limited opportunities to make changes to the local environment as they fall outside the powers of local partners, and where public health is not reflected in the local plan or planning not referenced in the health and wellbeing strategy or joint strategic needs assessment. Contexts in which the place-based working opportunities and synergies could be maximised were those where there were existing wider initiatives, regeneration activities or available development land that could be focused into the superzone.

#### Regional and national levers

The aims of the superzone pilot included an aspiration to use local areas’ experiences to identify, and change, levers at regional or national levels that could be used to facilitate local areas’ ambitions to create healthy zones around schools. However, these changes take time and this CMOC was unlikely to operate within a six month pilot period. There were lessons about raising expectations as superzone leads identified levers and were not aware of changes happening.

#### Measuring and demonstrating progress

A new CMOC that arose from the interviews related to the ability to measure and demonstrate progress to partners. The context for this was where local authority analytical capacity was available and baseline data was already available or could be collected. The outcome was continued partnership support for the project beyond the pilot phase.

## DISCUSSION

We conducted a process evaluation of the pilot using data collected from programme documents and interviews with the leads from all 13 superzones. The data from the document review provided an initial theory of the project, which we were then able to test with the data from the interviews. We identified 13 distinct context-mechanism-outcome configurations (CMOCs) which make up an initial programme theory of school superzones and which we present in a flow diagram (Figure 1). The results show that the internal partnership within the council and senior support were essential and highlight a range of other contexts and mechanisms that enhance a superzone. A strength of this study is that it offers the opportunity to observe 13 local authorities trying to implement the same broad model, helping to identify important contextual factors and mechanisms that affect how a superzone develops.

We found very little comparable research in the published literature, with regard to process evaluations of place-based partnership working to impact upon health and wellbeing through urban development. However, the importance of partnership working was clear in the findings of our research and we were able to locate some academic and grey literature on partnership working to improve health and wellbeing through urban regeneration schemes.[30–32]

Carley et al [30] highlight ‘foundation stones’ of partnerships, which can influence the quality of partnerships and their impact, but are outside of the control of the partnership. The establishment of the London devolution partnership could be viewed as one such ‘foundation stone’, with its role in enhancing collaboration between regional health and social care government partners, setting a coherent regional policy agenda for the development of superzones and providing system-wide support of the approach. The authors highlight that leadership at all levels of the partnership is crucial to the quality of partnerships, as is the competence of the partnership lead who may be required to nurture relationships with less enthusiastic partners at the beginning. The authors stress the need for a lead with the capacity to dedicate time to building and maintaining the partnership, a key finding in our research. Visioning and consensus building are vital components of effective partnerships, but so is the translation of the vision into deliverable actions. In contrast to our findings, the report by Carley et al[30] argues for the necessity of community involvement in partnerships, although this may reflect their focus on the impact of regeneration projects, rather than solely on process.

An academic review of the evidence base on the influence of partnership working in urban regeneration [31] highlights that the various agencies in partnerships differ in terms of their functions, their influence and their priorities. Partnership working should therefore be thought through and applied appropriately. This is consistent with our finding of the importance of the differing priorities and abilities of the partners to impact upon the environment of the superzone.

A 2009 review of the role of partnerships in changing the health and wellbeing of communities in urban regeneration areas proposes a ‘good enough’ approach to partnership working [32]. They argue that eight factors are needed to ensure a successful partnership, many of which echo the findings of this process evaluation: the right reasons, which are a shared vision and desire to work together; high stakes in the form of a strong need for partnership and contribution of finance and resources, as well as an agreed outcome; the right people with clear lines of accountability; the right leadership; strong relationships (which take time to develop); trust and respect; good communication; and formalising partnership arrangements (governance). Whilst the latter was not raised as a required element of a superzone, it does appear to be an element that would support the process and prevent some of the pitfalls, for example, the danger that a change in leadership or project lead undermines the superzone.

## Limitations

We do not present any data from those local authorities that chose not to be involved in the pilot, data that may add valuable insights into the contexts in which a superzone is unlikely to work and reasons why a local authority might choose not to be involved. The 13 pilot areas are also relatively homogeneous. Without variation in contextual factors, we are not able to say whether the lessons identified here would apply, for example in other urban contexts, or in councils with a different political make-up.

There are also important limits on the data that we present here on the 13 areas. The documents included in the initial document review only reflect the sponsoring organisations’ perspectives on the project. For each superzone, we only interviewed the superzone lead. Others involved in each superzone partnership may have different views about what happened and why. We were unable to identify published literature that report process evaluations of similar projects. This further limits our ability to say whether the data presented here reflects the experiences of similar projects elsewhere.

## CONCLUSION

This process evaluation represents a novel contribution to the evidence base on place-based approaches to urban environment and health. The limited research literature on this topic highlights the research value of this work and the potential for the full process and impact evaluation to add to the evidence base. While caution is needed in generalising from the experience of the pilot areas, the findings of this evaluation offer a basis for planning new superzones in areas that have not yet used this model, and for further testing an iteration of the programme theory.

## Data Availability

The data used are qualitative and to protect participants privacy, raw transcripts are not available

## RAMESES II CHECKLIST

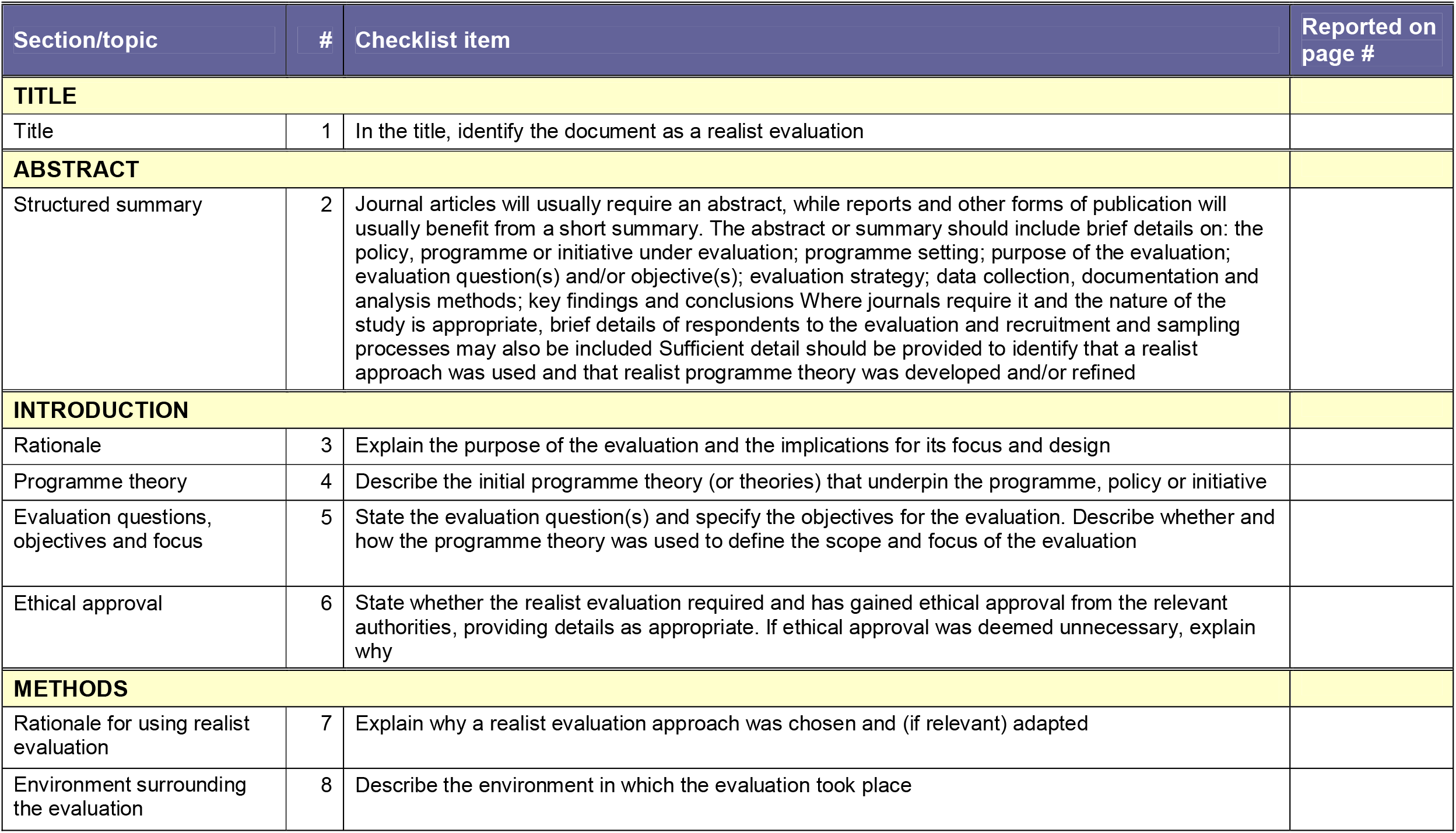
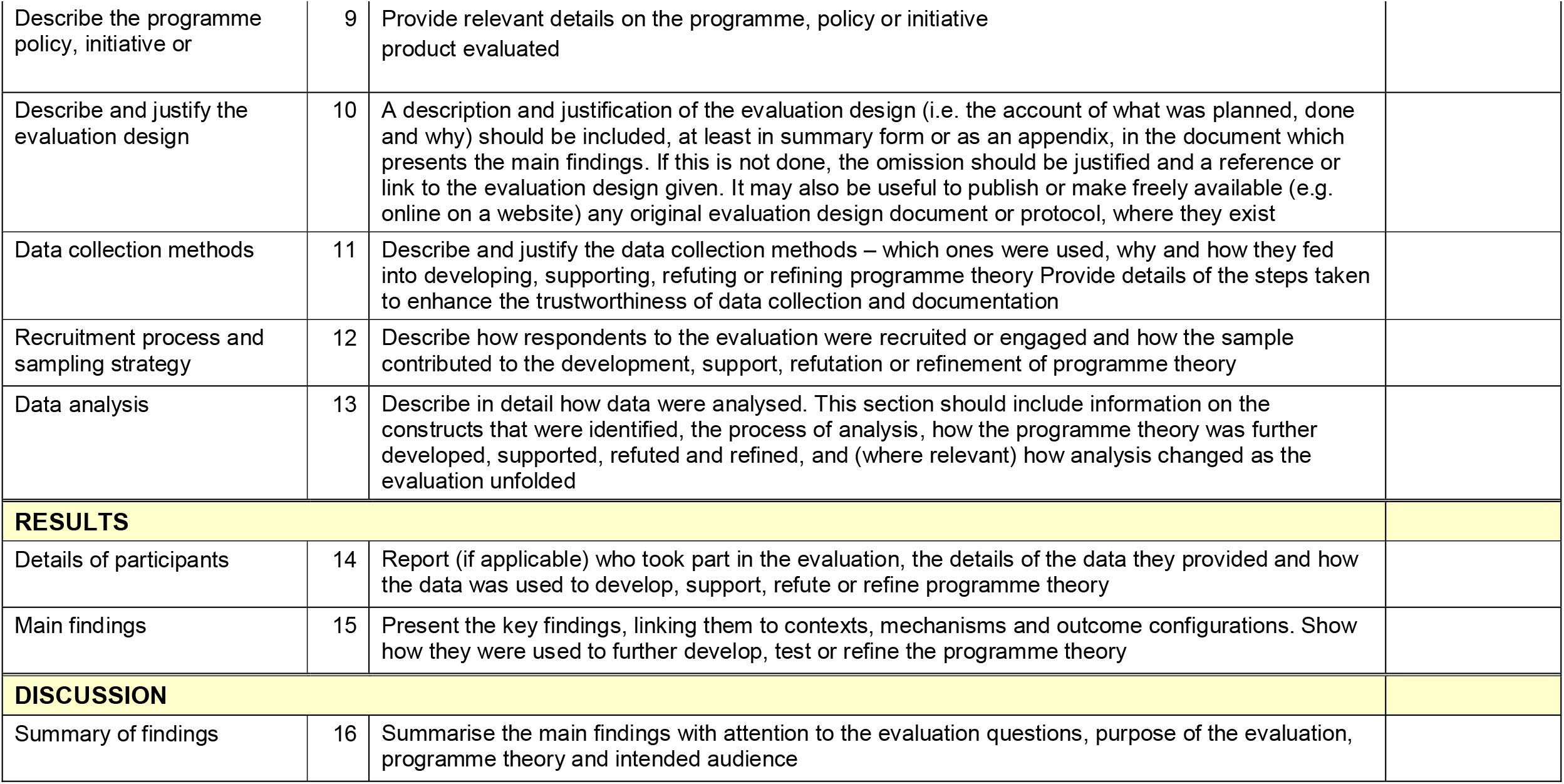
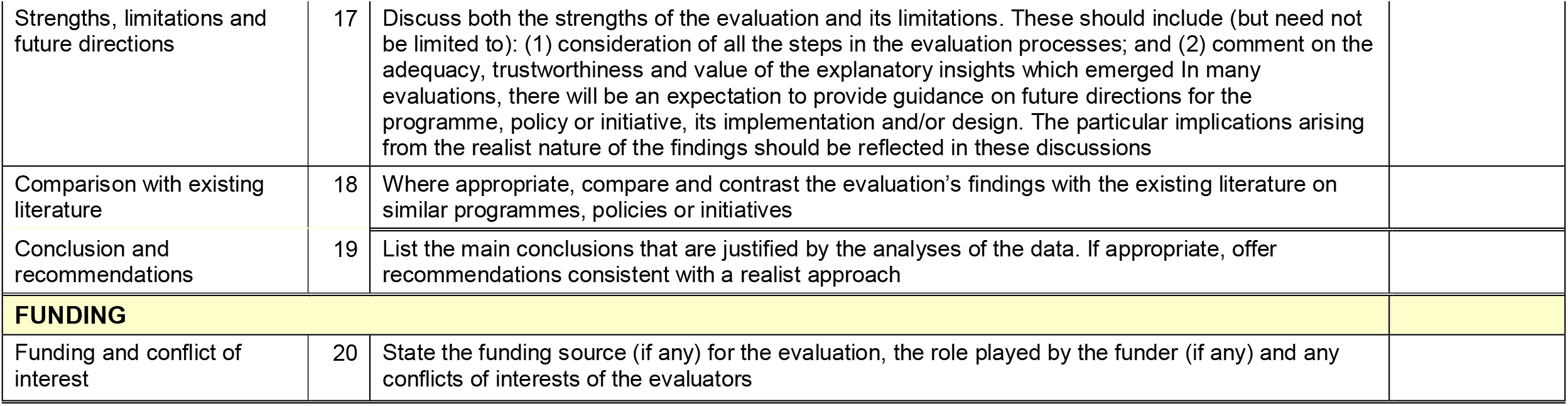

